# A Remote Household-Based Approach to Influenza Self-Testing and Antiviral Treatment

**DOI:** 10.1101/2021.02.01.21250973

**Authors:** Jessica Heimonen, Denise J. McCulloch, Jessica O’Hanlon, Ashley E. Kim, Anne Emanuels, Naomi Wilcox, Elisabeth Brandstetter, Mark Stewart, David McCune, Scott Fry, Sean Parsons, James P. Hughes, Michael L. Jackson, Timothy M. Uyeki, Michael Boeckh, Lea M. Starita, Trevor Bedford, Janet A. Englund, Helen Y. Chu, on behalf of Seattle Flu Study investigators

## Abstract

**Background:** Households represent important settings for transmission of influenza and other respiratory viruses. Current influenza diagnosis and treatment relies upon patient visits to healthcare facilities, which may lead to under-diagnosis and treatment delays. This study aimed to assess the feasibility of an at-home approach to influenza diagnosis and treatment via home testing, telehealth care, and rapid antiviral home delivery.

**Methods:** We conducted a pilot interventional study of remote influenza diagnosis and treatment in Seattle-area households with children during the 2019-2020 influenza season using pre-positioned nasal swabs and home influenza tests. Home monitoring for respiratory symptoms occurred weekly; if symptoms were reported within 48 hours of onset, participants collected mid-nasal swabs and used a rapid home-based influenza immunoassay. An additional home-collected swab was returned to a laboratory for confirmatory influenza RT-PCR testing. Baloxavir antiviral treatment was prescribed and delivered to symptomatic and age-eligible participants, following a telehealth encounter.

**Results:** 124 households comprising 481 individuals self-monitored for respiratory symptoms, with 58 home tests administered. 12 home tests were positive for influenza, of which 8 were true positives confirmed by RT-PCR. The sensitivity and specificity of the home influenza test was 72.7% and 96.2%, respectively. There were 8 home deliveries of baloxavir, with 7 (87.5%) occurring within 3 hours of prescription, and all within 48 hours of symptom onset.

**Conclusions:** We demonstrate the feasibility of self-testing combined with rapid home delivery of influenza antiviral treatment. This approach may be an important control strategy for influenza epidemics and pandemics.

**Summary:** In this pilot study, 481 individuals self-monitored for respiratory symptoms. Of 58 home tests, 12 were influenza-positive. There were 8 baloxavir home deliveries within 48 hours of illness onset. A home-based approach to influenza diagnosis and treatment could be feasible.

## Introduction

In the United States, influenza is typically diagnosed during an in-person healthcare visit and if antiviral treatment is prescribed, a subsequent visit to a pharmacy is required. This multi-step process may lead to delays in receipt of antivirals and potentially exposes other individuals in clinics and pharmacies to influenza. Since antiviral therapy is most effective when started within 48 hours of symptom onset, reducing delays to treatment initiation may improve outcomes in treated persons.^1-3^ Baloxavir is an oral FDA-approved antiviral for early treatment of uncomplicated influenza in individuals aged 12 years and older. The long half-life of baloxavir allows a single treatment dose in contrast to five twice-daily doses of oseltamivir. Moreover, baloxavir treatment is associated with shorter duration of influenza virus detection compared with oseltamivir or placebo.^4^

Households, particularly those with young children, play a key role in seasonal influenza epidemics because the frequency and intensity of contacts among household members are greater than in the broader community.^5^ Prior studies have shown that young children are important contributors to the introduction and transmission of influenza in households. ^6,7^ Therefore, households represent an important setting to study influenza intervention strategies.

Home-based influenza testing and rapid treatment with home-delivered antivirals have not been evaluated in clinical trials. Home diagnosis of respiratory infections via self-testing or telemedicine services has the potential for widespread use, particularly during a pandemic where periods of social distancing and restricted movement occur. Similarly, home-based initiation of antiviral therapy may decrease time from symptom onset to initiation of therapy and could improve outcomes compared with current management practices. Advances in telemedicine services (telehealth), rapid delivery services, and the ongoing development of home-based influenza assays may make this a feasible strategy to employ. Here we report the results of a pilot study examining the feasibility of a test-and-treat method for influenza in households with children, including the use of home influenza testing, telehealth, and rapid antiviral delivery.

## Methods

### Study Design

We conducted a prospective interventional study to assess the feasibility of a home-based approach to diagnosis and treatment of influenza in households with children. This study was conducted in the Seattle metropolitan area as part of the Seattle Flu Study.^8^ The recruitment process and eligibility criteria were previously described.^9^ Briefly, households of ≥3 individuals sleeping in the home for ≥4 days per week, with at least one child aged three months to 17 years, and containing ≥2 baloxavir age-eligible individuals, were eligible to participate. Recruitment was conducted via web-based advertisements and social media. Households were consented, and all data were captured using a remote, electronic platform in Project REDCap (Research Electronic Data Capture).^10^ All informed consent conferences took place via phone, with written consent by household members.

At enrollment, one household member was designated the lead contact and provided demographic and baseline health information about all household members. All enrolled households were asked to complete a weekly survey regarding the presence or absence of acute respiratory infection (ARI) symptoms. ARI was defined as new or worsening acute cough or the presence of two or more respiratory symptoms (**Table A1**). Recruitment started in November 2019, and beginning on December 23, 2019, individuals self-reporting ARI within 48 hours of symptom onset self-collected or had a parent collect two mid-nasal swabs (Copan, Murrieta, CA): one to perform a rapid home-based influenza immunoassay (Ellume, East Brisbane, Queensland, Au), and one for confirmatory reverse transcription polymerase chain reaction (RT-PCR) testing. Individuals reporting ARI and with a positive home influenza test result were linked to telehealth care (98point6, Seattle, WA) if eligible for baloxavir (age ≥12 years and otherwise healthy or at increased risk of developing influenza-related complications, excluding individuals with cancer, immunosuppression, liver or kidney disease). If a diagnosis of influenza was supported by a telehealth provider’s review of the patient’s symptoms, in addition to the positive home influenza test result, a prescription for baloxavir was sent to the study pharmacy (University of Washington [UW] Investigational Drug Service, Seattle, WA). Following dispensing, baloxavir was delivered to the household via a rapid courier service (Delivery Express, Tukwila, WA and FedEx, Memphis, TN) scheduled remotely by the study team. One week after swab collection, ill participants were asked to complete a follow-up questionnaire reporting illness outcomes, the usability of the home test, as well as hypothetical illness behavioral changes with and without use of the home influenza test.

On February 7, 2020, there was a modification to the study design due to a required protocol change that prohibited the return of the influenza home test results to participants and telehealth providers. Thus, the data presented here reflect the study period up until February 7, 2019 only. This study is registered on ClinicalTrials.gov (NCT04141930) and was approved by the UW Institutional Review Board.

### Rapid Home Influenza Testing

The rapid home-based influenza immunoassay was developed, produced, and manufactured by Ellume (Ellume, East Brisbane, Queensland, Au). This antigen detection test uses a combination of bioluminescence and Bluetooth technology, where users self-collect a mid-nasal swab and then use device-specific equipment to add their sample to an analyzer. The analyzer conducts the rapid assay, testing against influenza A and influenza B virus targets, then sends the result to a participant’s smartphone using Bluetooth.

### Laboratory Testing

Home-collected nasal specimens were placed in universal transport media (UTM) (Becton, Dickinson and Company, Sparks, MD) in accordance with International Air Transport Association (IATA) guidelines and transported to the laboratory at ambient temperature generally within 48-72 hours of collection where samples were aliquoted at room temperature and stored at 4°C prior to testing. Samples were extracted (Magnapure 96, Roche, Basel, CH) and tested for respiratory pathogens, including influenza virus types and influenza A subtypes, by TaqMan RT-PCR (Thermofisher, Waltham, MA) on a QuantStudio 12 (Applied Biosystems, Foster City, CA) (**Table 2A)**. Positive and negative controls were included in each extraction and RT-PCR run. All samples were tested for Rnase P, a human cellular marker, and Rnase P relative cycle threshold (Crt) values were used to evaluate sample quality.

**Table 2.**
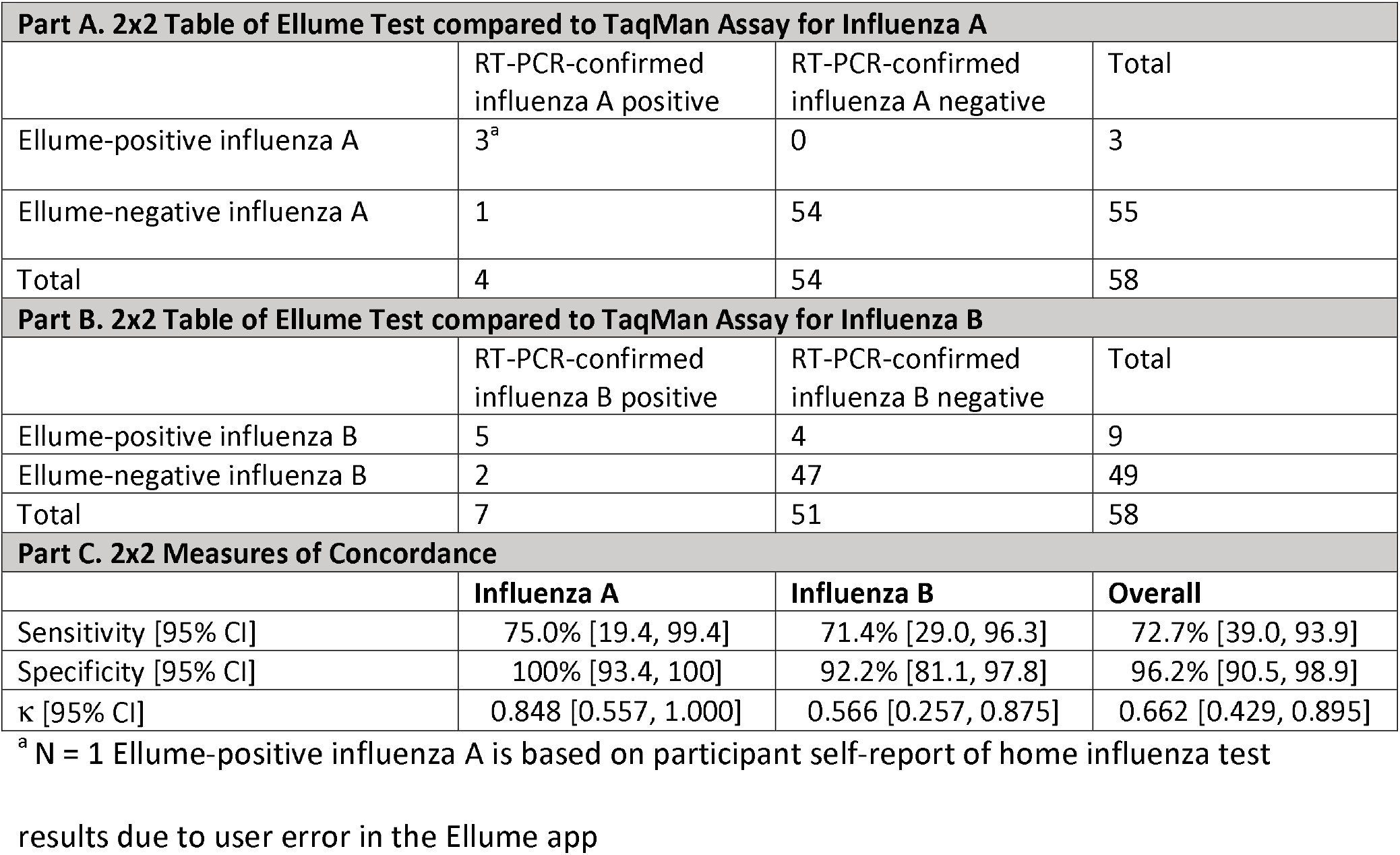
Home influenza test results in comparison to RT-PCR-confirmed influenza test results

| **Part A. 2x2 Table of Ellume Test compared to TaqMan Assay for Influenza A** |  |  |  |
| --- | --- | --- | --- |
|  | RT-PCR-confirmed influenza A positive | RT-PCR-confirmed influenza A negative | Total |
| Ellume-positive influenza A | 3<sup>a</sup> | 0 | 3 |
| Ellume-negative influenza A | 1 | 54 | 55 |
| Total | 4 | 54 | 58 |
| **Part B. 2x2 Table of Ellume Test compared to TaqMan Assay for Influenza B** |  |  |  |
|  | RT-PCR-confirmed influenza B positive | RT-PCR-confirmed influenza B negative | Total |
| Ellume-positive influenza B | 5 | 4 | 9 |
| Ellume-negative influenza B | 2 | 47 | 49 |
| Total | 7 | 51 | 58 |
| **Part C. 2x2 Measures of Concordance** |  |  |  |
|  | Influenza A | Influenza B | Overall |
| Sensitivity [95% CI] | 75.0% [19.4, 99.4] | 71.4% [29.0, 96.3] | 72.7% [39.0, 93.9] |
| Specificity [95% CI] | 100% [93.4, 100] | 92.2% [81.1, 97.8] | 96.2% [90.5, 98.9] |
| $\kappa$ [95% CI] | 0.848 [0.557, 1.000] | 0.566 [0.257, 0.875] | 0.662 [0.429, 0.895] |

<sup>a</sup> N = 1 Ellume-positive influenza A is based on participant self-report of home influenza test
results due to user error in the Ellume app

### Statistical Analysis

Analyses were restricted to enrolled households that completed at least one symptom log prior to February 7, 2020. The illness results presented here are confined to specimens that were collected and received in the laboratory by February 7, 2020. Participant-level demographic information is reported by RT-PCR-confirmed influenza status. Chronic respiratory disease was defined as a history of asthma or reactive airway disease, COPD or emphysema, or chronic bronchitis. Other chronic diseases such as diabetes, heart failure, or cancer were defined as non-respiratory chronic disease. Participant-reported home influenza test usability, hypothetical behavioral changes when ill with and without the use of the home influenza test, as well as the sensitivity, specificity, and Cohen’s kappa coefficient (κ) were calculated; concordance measures compared the influenza home test with the TaqMan assay, where the TaqMan assay represented the gold standard. A p-value <0.05 was considered statistically significant. All analyses were conducted using SAS software version 9.4.

## Results

### Demographics

From November 2019 to February 7, 2020, 150 households enrolled in the study; 124 households completed one or more weekly symptom logs, resulting in 481 unique individuals self-monitoring for respiratory symptoms (**Figure 1B**). Overall, the study population was mostly comprised of healthy individuals, with 89.9% of the population reporting no chronic health conditions (**Table 1**). Most individuals were insured, and 79.4% reported receiving the seasonal influenza vaccine. The study population predominately consisted of white individuals aged 18 to 49 years (44.7%) or 5 to 17 years (37.4%).

**Table 1.** Participant demographics characteristics from enrolled households by RT-PCR-confirmed influenza status based on influenza cases detected during the Test-and-Treat Strategy

| **Participant Demographics** | **Row Total** | **Influenza-positive** | **Influenza-negative** |
| --- | --- | --- | --- |
| *N* | 481 | 11 | 470 |
| *Age* |  |  |  |
| 0-4 years <sup>a</sup> | 41 | 2 (4.9) | 39 (95.1) |
| 5-17 years | 180 | 4 (2.2) | 176 (97.8) |
| 18-49 years | 215 | 5 (2.3) | 210 (97.7) |
| 50-64 years | 35 | 0 (0.0) | 35 (100) |
| ≥65 years | 10 | 0 (0.0) | 10 (100) |
| *Female Sex* | 253 | 8 (3.2) | 245 (96.8) |
| *Race* <sup>b</sup> |  |  |  |
| Asian | 9 | 0 (0.0) | 9 (100) |
| Black | 8 | 8 (0.0) | 8 (100) |
| White | 408 | 7 (1.7) | 401 (98.3) |
| Other | 14 | 1 (7.1) | 13 (92.9) |
| Multiple | 31 | 3 (9.7) | 28 (90.3) |
| *Hispanic/Latino* <sup>c</sup> | 35 | 2 (5.7) | 33 (94.3) |
| *High-risk condition* <sup>d</sup> |  |  |  |
| Respiratory | 50 | 2 (4.0) | 48 (96.0) |
| Other | 16 | 0 (0.0) | 16 (100) |
| None | 412 | 8 (1.9) | 404 (98.1) |
| *Insured* | 480 | 11 (2.3) | 469 (97.7) |
| *Received Influenza Vaccine* <sup>e</sup> | 382 | 10 (2.6) | 372 (97.4) |

<sup>a</sup> Includes those aged 6 months and older
<sup>b</sup> N = 10 individuals were missing information about race
<sup>c</sup> N = 7 individuals were missing information about ethnicity
<sup>d</sup> N = 3 individuals missing information about medical history
<sup>e</sup> N = 1 individuals missing information on influenza vaccination status

**Figure 1A.**
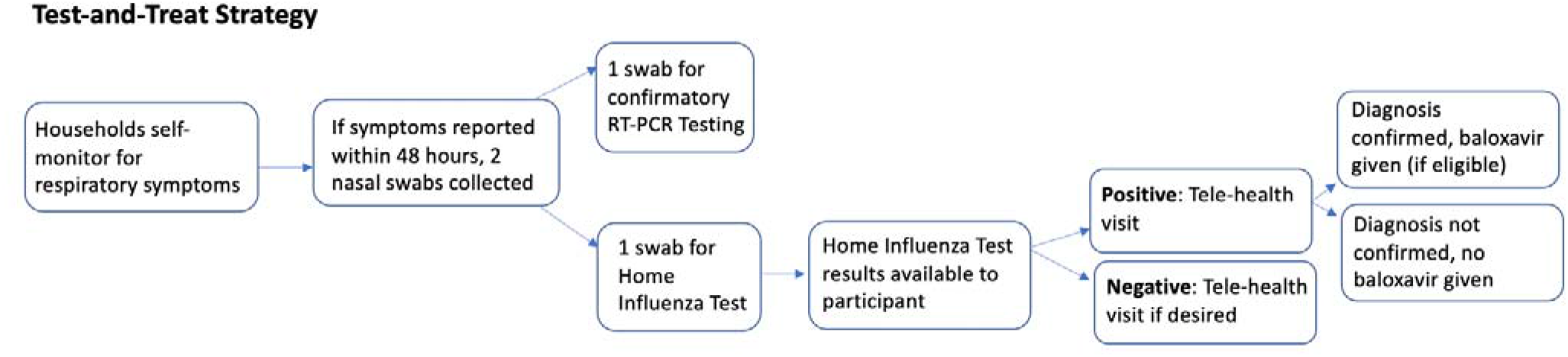
Study design overview including household-level and participant-level study flow of the Test-and-Trest Strategy from December 23, 2019, to Feburary 7, 2020.

**Figure 1B.**
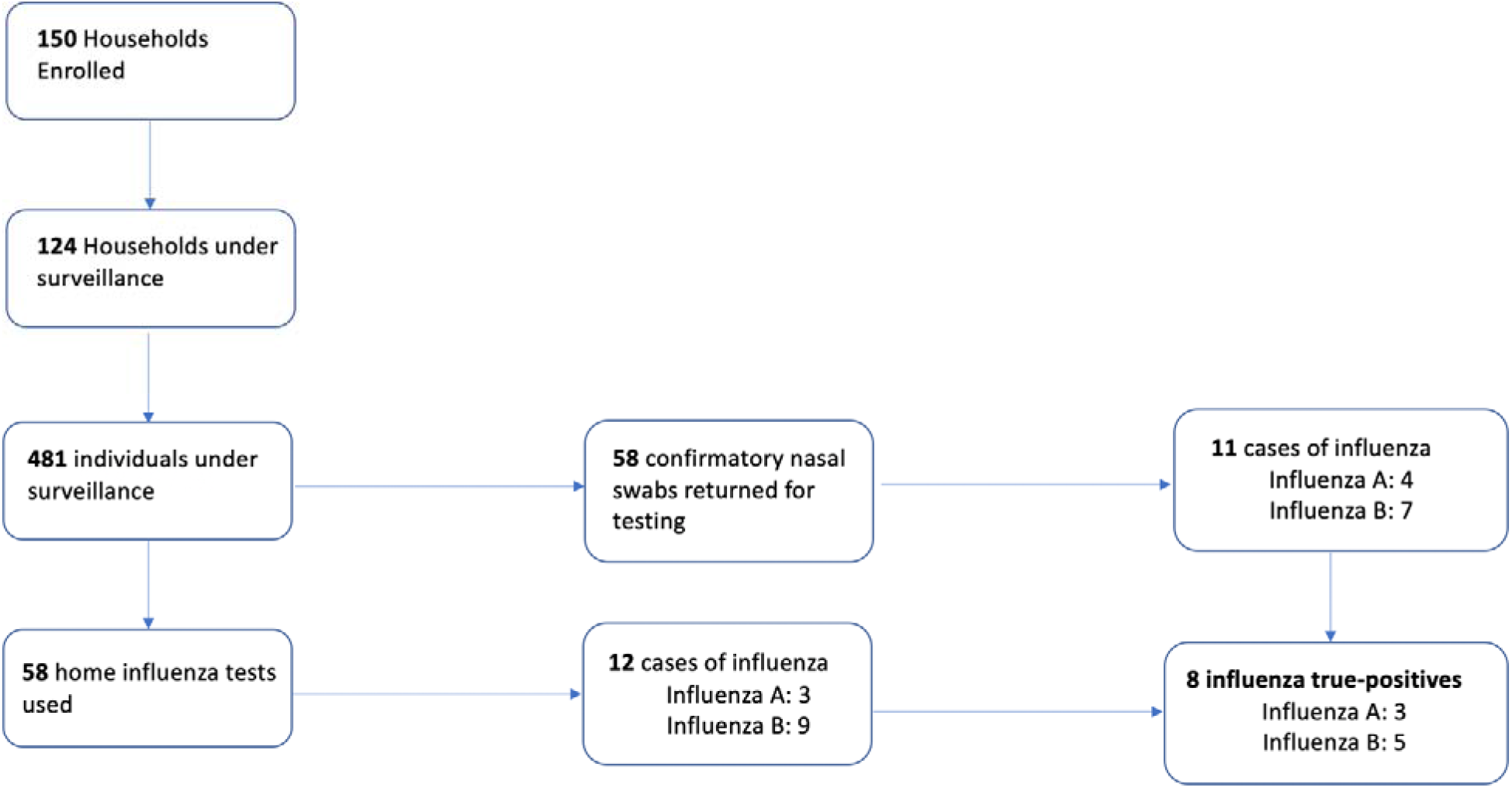
Total number of households and particpants completing study procedures steps based on households initiating symptom monitoring prior to February 7, 2020.

### Home Influenza Self-Testing

Among participants experiencing respiratory symptoms, 58 influenza home tests were used during the test-and-treat phase of the study, yielding 12 positive results. Home influenza test results were compared with RT-PCR results from the confirmatory nasal swabs (**Table 2**).

Measures of agreement of the home influenza test were similar for influenza A and influenza B, though measures of agreement were higher for influenza A than influenza B: 75.0% sensitivity and 100% specificity for influenza A, 71.4% sensitivity and 92.2% specificity for influenza B. Likewise, Cohen’s kappa was higher for influenza A (κ = 0.848) compared with influenza B (κ = 0.566). Notably, the majority of false positives were influenza B, while the percent of false negatives were similar for influenza A (25%) and influenza B (28.5%). The overall sensitivity of the home influenza test was 72.7% and the specificity was 96.2%, suggesting home test performance was concordant with RT-PCR (κ = 0.633).

Among 47 participants who used the home influenza test and completed the follow-up questionnaire, 93.6% reported that experiencing respiratory symptoms and a positive result would lead to minimizing contact with others while 89.4% reported that experiencing respiratory symptoms and a positive result would lead to missing work or school (**Table 3**). In contrast, 78.7% reported they would minimize contact with others if experiencing respiratory symptoms but no home test result or diagnosis was available, while 59.6% reported they would miss work or school if experiencing respiratory symptoms but no home test result or diagnosis was available.

**Table 3.** Home influenza test usability findings from participants that used a home influenza test and completed the one-week follow-up illness questionnaire

| **Home Flu Test Self-Reported Participant Feedback (N = 47)** |  | **N (%)** |  |
| --- | --- | --- | --- |
| Miss school or work if having respiratory symptoms and test was positive |  | 42 (89.4) |  |
| Miss school or work if having respiratory symptoms but no test was used or diagnosis provided |  | 28 (59.6) |  |
| Minimize contact with others if having respiratory symptoms and test was positive |  | 44 (93.6) |  |
| Minimize contact with others if having respiratory symptoms but no test was used or diagnosis provided |  | 37 (78.7) |  |
|  | **Agree (%)** | **Neutral (%)** | **Disagree (%)** |
| Test was easy to use (N = 46) | 39 (83.0) | 3 (6.4) | 4 (8.5) |
| App was easy to use (N = 46) | 42 (89.4) | 2 (4.3) | 2 (4.3) |
| Results were easy to read (N = 46) | 40 (85.1) | 4 (8.5) | 2 (4.3) |
| Felt confident using the test (N = 46) | 40 (85.1) | 4 (8.5) | 2 (4.3) |

### Telehealth Influenza Diagnosis and Treatment

Among participants experiencing ARI, there were 11 telehealth visits (**Table 4**). In total, there were 8 baloxavir home deliveries. The median delivery time was 1.62 hours; 87.5% of home deliveries occurred within 3 hours from the time of baloxavir prescription. Twenty-five percent of deliveries occurred within 24 hours of symptom onset, 37.5% occurred within 30 hours of symptom onset, and 37.5% occurred 30 to 48 hours after symptom onset.

**Table 4.**
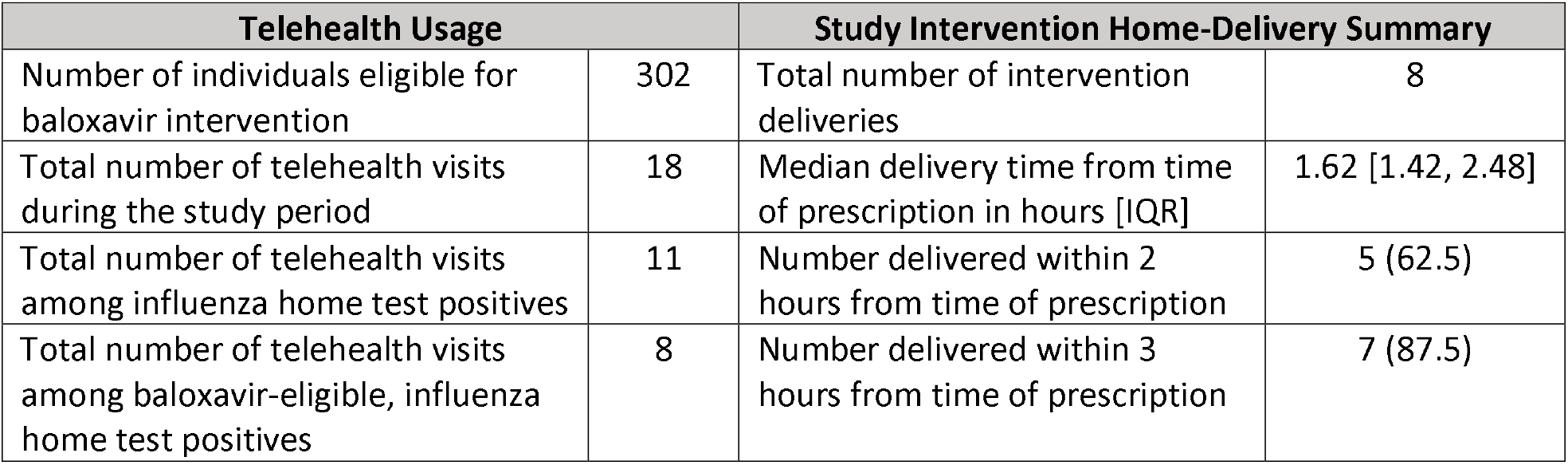
Participant telehealth usage and home antiviral delivery results from December 23, 2019 to February 7, 2020

| Telehealth Usage |  | Study Intervention Home-Delivery Summary |  |
| --- | --- | --- | --- |
| Number of individuals eligible for baloxavir intervention | 302 | Total number of intervention deliveries | 8 |
| Total number of telehealth visits during the study period | 18 | Median delivery time from time of prescription in hours [IQR] | 1.62 [1.42, 2.48] |
| Total number of telehealth visits among influenza home test positives | 11 | Number delivered within 2 hours from time of prescription | 5 (62.5) |
| Total number of telehealth visits among baloxavir-eligible, influenza home test positives | 8 | Number delivered within 3 hours from time of prescription | 7 (87.5) |

### Laboratory Testing

There were 58 nasal swabs collected concurrently with the home influenza test at the time of illness. These were returned to the laboratory for RT-PCR, and yielded 11 (19.0%) influenza-positive cases from 11 individuals (7 influenza B and 4 influenza A) (**Figure 1B**). Among influenza cases, 4 were baloxavir-ineligible due to age or medical history and 7 were eligible, but 3 of these 7 baloxavir-eligible influenza cases were not treated. Two of these 3 (66.7%) individuals had false negative results by home influenza test compared with RT-PCR, and 1 (33.3%) opted not to pursue telehealth care. Likewise, 4 influenza-negative individuals received antiviral baloxavir treatment; all of these individuals had false positive home test results compared with RT-PCR. Overall, there were 4 RT-PCR-confirmed influenza-positive and 4 RT-PCR-confirmed influenza-negative individuals who received baloxavir.

## Discussion

To our knowledge, this is the first report of a remote, household-based approach to influenza diagnosis and treatment in which no face-to-face contact with a healthcare provider or pharmacist was required. In this pilot study, participants successfully self-monitored for the onset of respiratory symptoms, self-conducted a rapid home influenza test, remotely discussed their illness with a healthcare provider, and received prompt delivery of a prescribed antiviral medication when indicated.

Participants were adherent to study procedures, with 124 (82.6%) of households participating in weekly respiratory surveillance and 58 successfully completing home influenza tests. The vast majority of participants reported that the home influenza test and app were easy to use and that the results were easy to understand.

The results of rapid home influenza testing were largely concordant with RT-PCR. Concordance was higher for influenza A than influenza B. Notably, the test-and-treat strategy encompassed only a part of the 2019-2020 influenza season. In particular, cases of influenza B predominantly occurred prior to influenza A cases, which is unusual but consistent with other results published for the 2019-2020 influenza season.^14^ Thus, the measures of concordance for influenza B may be skewed due to the timeline of when the test-and-treat strategy started.

Home influenza test results may have assisted telehealth providers in making an accurate influenza diagnosis. Previous studies have demonstrated that influenza diagnosis based on a provider review of symptoms has low sensitivity; ^15,16^ adding a sensitive home-based test has the potential to significantly improve influenza diagnostic accuracy.

A small number of influenza-positive participants received baloxavir, yet our results suggest a home-delivery approach is feasible, particularly because all 8 individuals received drug within 48 hours of symptom onset, and 87.5% of home-deliveries arrived within 3 hours from the time of prescription. Four influenza-negative individuals received baloxavir therapy, although no adverse effects were observed and no major differences were seen among baloxavir treated and untreated groups.

Our remote approach to home testing and treatment of influenza may be an important future control strategy, particularly during a severe epidemic or pandemic,^17^ and even with non-influenza viruses, such as SARS-CoV-2. Current reports suggest a version of this strategy may be operational for the 2020-2021 influenza season.^18^ The potential public health importance of a home-based test and treat strategy is supported by the large percentage of participants who reported that a positive influenza test result would influence their behavior, such as limiting contact with others or not attending work or school while sick, compared with the reported lack of behavioral change from experiencing respiratory symptoms without any test result or diagnosis. Moreover, the majority of our participants received baloxavir antiviral treatment within 30 hours of symptom onset. While baloxavir confers the greatest clinical benefit when initiated within 24 hours^4^, our findings suggest that rapid antiviral-home delivery is feasible and that a remote approach to influenza diagnosis and treatment can decrease the time from symptom onset to initiation of antiviral therapy.

There are several limitations to this study. First, the results of this pilot study encompassed only part of one influenza season, and thus did not capture the peak of local influenza A virus transmission, leading to a small sample size for that pathogen. Data were based on self-collection and self-report, which may be subjective particularly for variables such as symptoms or illness duration. Furthermore, despite good compliance with study procedures, there were a few instances of participants collecting nasal swabs without providing clinical information. Additionally, these results were derived from a largely homogeneous volunteer study population of highly educated, middle to upper-class, white households, and may limit the generalizability of the results. The results presented here are also limited by the antiviral therapy being prescribed to a small number of households in a regulated, well-resourced study environment. Further studies are needed to assess the feasibility of this home-based influenza test and rapid home antiviral delivery strategy in larger or more remote populations.

## Conclusions

The moderate sensitivity of the rapid home influenza test coupled with successful antiviral home-delivery suggest that the implementation of intervention or control strategies in households with children could be feasible and may be particularly useful when circumstances dictate restricted movement or social distancing. Further studies on this topic would help understand the usefulness of these strategies in more remote or diverse populations. While the strategy for early diagnosis and treatment of influenza was studied, it has the potential to be applied to other respiratory viruses that cause epidemics and pandemics as home-based diagnostic and treatment options become available.

## Data Availability

Data and code used for analyses may be available upon request.

## Funding

The Seattle Flu Study is funded by Gates Ventures. The funder was not involved in the design of the study, does not have any ownership over the management and conduct of the study, the data, or the rights to publish.

## Conflicts of Interest

H.Y.C. has received research support from GlaxoSmithKline, Novavax, and Sanofi Pasteur; J.A.E. has received research support from AstraZeneca, GlaxoSmithKine, Merck, and Pfizer and served as a consultant for Sanofi Pasteur and Meissa Vaccines. M.L.J. has received research support from Sanofi Pasteur. M.B. receives research support and serves as a consultant for Ansun Biopharma, Gilead Sciences, Janssen, and Vir Biotechnology; and serves as a consultant to GlaxoSmithKline, ReViral, ADMA, Pulmocdie and Moderna. All other authors: J.H, D.J.M, J.O, A.E.K, A.E, N.W, E.B, M.S, D.M, S.F, S.P, J.P.H, L.M.S, T.B, T.M.U. have no conflicts to report.

## Acknowledgements

We gratefully acknowledge the Seattle Flu Study team for their work on this project, as well as the University of Washington Montlake Investigational Drug Service and Inpatient pharmacies for serving as our 24/7 study pharmacy. Thank you to the 98point6 team of providers that helped make this project functional. We would like to thank the participating households for the time they spent conducting our study procedures. REDCap at ITHS is supported by the National Center for Advancing Translational Sciences of the National Institutes of Health under Award Number UL1 TR002319.

## Supplemental Tables

**Table A1.**
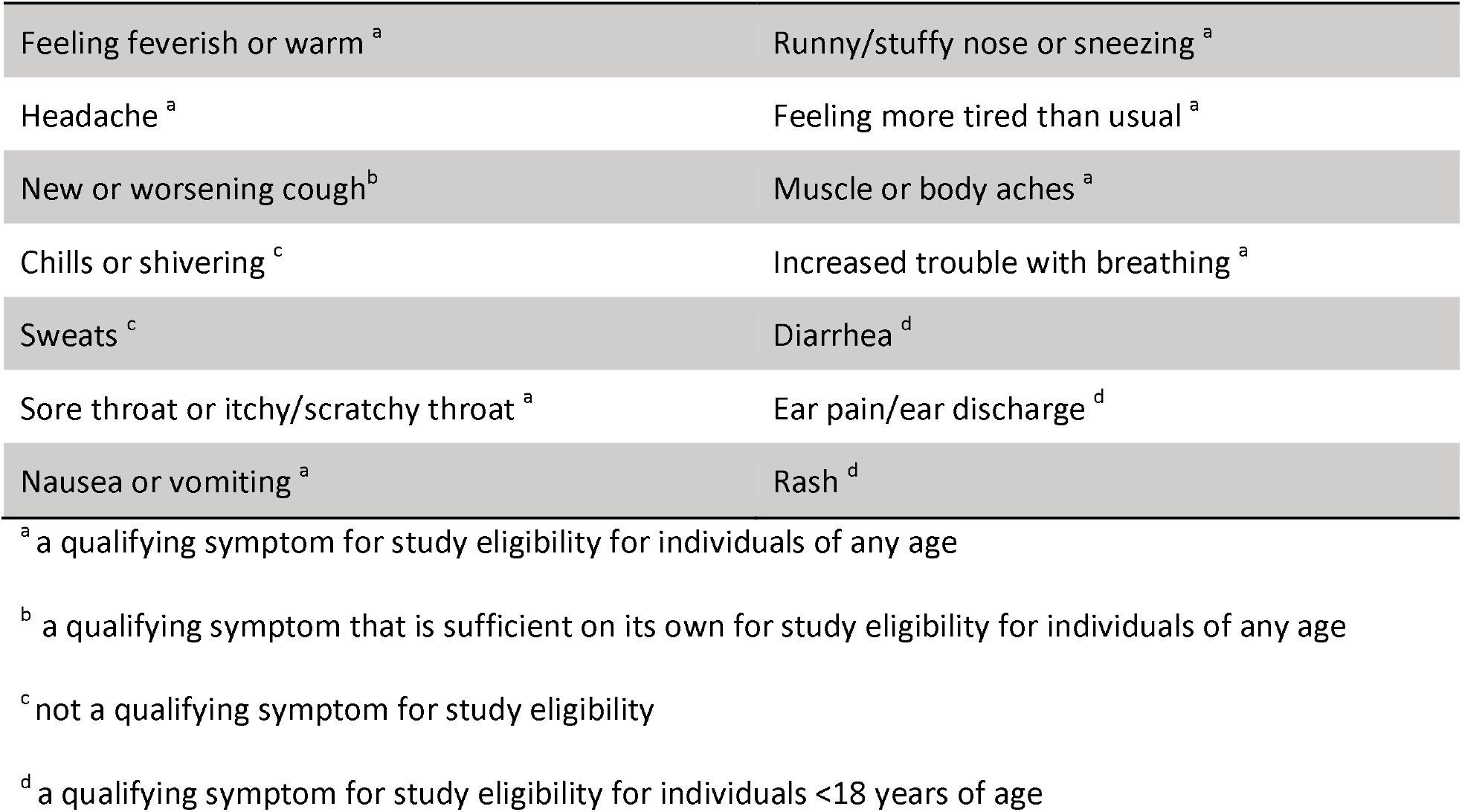
List of symptoms used to determine for eligibility for nasal swab collection. Acute cough or two or more concurrent *qualifying* symptoms was considered an acute illness episode

|  |  |
| --- | --- |
| Feeling feverish or warm <sup>a</sup> | Runny/stuffy nose or sneezing <sup>a</sup> |
| Headache <sup>a</sup> | Feeling more tired than usual <sup>a</sup> |
| New or worsening cough <sup>b</sup> | Muscle or body aches <sup>a</sup> |
| Chills or shivering <sup>c</sup> | Increased trouble with breathing <sup>a</sup> |
| Sweats <sup>c</sup> | Diarrhea <sup>d</sup> |
| Sore throat or itchy/scratchy throat <sup>a</sup> | Ear pain/ear discharge <sup>d</sup> |
| Nausea or vomiting <sup>a</sup> | Rash <sup>d</sup> |

<sup>a</sup> a qualifying symptom for study eligibility for individuals of any age
<sup>b</sup> a qualifying symptom that is sufficient on its own for study eligibility for individuals of any age
<sup>c</sup> not a qualifying symptom for study eligibility
<sup>d</sup> a qualifying symptom for study eligibility for individuals <18 years of age

**Table A2.** Pathogens for which respiratory specimens are tested using a TaqMan RT-PCR.

| Viruses | Bacteria |
| --- | --- |
| Influenza A/H1N1 | *Streptococcus pneumoniae* |
| Influenza A/H3N2 | *Mycoplasma pneumoniae* |
| Influenza B | *Chlamydia pneumoniae* |
| Influenza C |  |
| Respiratory syncytial viruses (RSV) A and B |  |
| Parainfluenza viruses 1-4 |  |
| Human coronaviruses 229E, NL63, OC43, and HKU1 |  |
| Adenovirus |  |
| Human rhinovirus |  |
| Measles |  |
| Mumps |  |
| Human metapneumovirus |  |
| Human parechovirus |  |
| Enterovirus, non-D-68 <sup>a</sup> |  |
| Enterovirus D68 |  |
| Human bocavirus |  |

<sup>a</sup> All enterovirus species A, B, C, D, and G, including: all Coxsackie serotypes under species A, B, C; all
Echovirus serotypes; all Poliovirus serotypes (1-3)

## Notes

### Clinical Trial

NCT04141930

### Author Declarations

University of Washington Institutional Review Board (STUDY00008200)

